# How to incentivise doctor attendance in Bangladesh: a latent class analysis of a discrete choice experiment

**DOI:** 10.1101/2021.03.21.21254042

**Authors:** Blake Angell, Mushtaq Khan, Mir Raihanul Islam, Kate Mandeville, Nahitun Naher, Eleanor Hutchinson, Martin McKee, Syed Masud Ahmed, Dina Balabanova

**Author notes:** Corresponding author. Level 5, 1 King Street, Newtown, New South Wales, Australia. **Funding:** This publication is an output of the SOAS Anti-Corruption Evidence (ACE) research consortium funded by UK aid from the UK Government [Contract P07073]. The views presented are those of the author(s) and do not necessarily reflect the UK government’s official policies or the views of SOAS-ACE or other partner organisations. For more information on SOAS-ACE visit www.ace.soas.ac.uk.

## Abstract

**Objective:** To elicit preferences of doctors over interventions to address doctor absenteeism in rural facilities in Bangladesh, a pervasive form of corruption across the country.

**Methods:** We conducted a discrete choice experiment with 308 doctors across four tertiary hospitals in Dhaka, Bangladesh. Four attributes of rural postings were included based on a literature review, qualitative research and a consensus-building workshop with policymakers and key health-system stakeholders: relationship with the community, security measures, attendance-based policies, and incentive payments. Respondents’ choices were analysed with mixed multinomial logistic and latent class models and were used to simulate the likely uptake of jobs under different policy packages.

**Results:** All attributes significantly impacted doctor choices (p<0.01). Doctors strongly preferred jobs at rural facilities where there was a supportive relationship with the community (β=0.93), considered good attendance in education and training (0.77) or promotion decisions (0.67), with functional security (0.67) and higher incentive payments (0.5 per 10% increase of base salary). Jobs with disciplinary action for poor attendance were disliked by respondents (-.63). Latent class analysis identified three groups of doctors that differed in their uptake of jobs. Scenario modelling identified intervention packages that differentially impacted doctor behaciour and combinations that could feasibly improve doctors’ attendance.

**Conclusion:** Bangladeshi doctors have strong but varied preferences over interventions to overcome absenteeism. Some were unresponsive to intervention but a substantial number appear amenable to change. Designing policy packages that consider these differences and target particular doctors could begin to generate sustainable solutions to doctor absenteeism in rural Bangladesh.

## INTRODUCTION

Health worker absenteeism is widespread in many low- and middle-income countries and is a major obstacle to achieving universal health coverage as it disproportionately affects vulnerable groups [1-3]. In Bangladesh, newly appointed doctors must serve at least two years in rural areas and at lower administrative tiers of government (reduced to one year for a few remote areas) [4, 5]. Absenteeism from these facilities is a violation of their employment contracts and can be considered a form of corruption. In Bangladesh it is estimated that 40% of doctors are absent at any time, rising to 74% at facilities staffed by a single doctor, typically in rural areas covering already underserved communities [3]. Traditionally, absenteeism has been viewed as resulting from ineffective oversight, weak governance, and inadequate enforcement, so interventions have focused on top-down measures to enforce compliance. For example, the government installed fingerprint scanners to verify attendance by health workers in rural health clinics, publishing attendance rates online [6]. This has had little effect, however, as with similar strategies used elsewhere in Bangladesh and beyond [7-10].

Recent work has recast absenteeism as a structural rather than moral issue [11], with increased recognition of broader systemic factors driving doctor absenteeism. These include poor working conditions, misaligned incentives for doctors and those monitoring attendance, security of rural facilities and preferences to live in urban rather than the rural locations where absenteeism is particularly acute [4, 12, 13]. The political settlements analysis developed by Khan and others [14, 15] identifies the distribution of power and capabilities across organisations as important explanators of rent-seeking and rule-violating behaviours that affect the effectiveness of enforcement strategies. Failure to enforce doctors’ attendance where the rule-of-law is weak may be explained by their organisational power and the many ‘reasonable’ reasons why many doctors are absent. Enforcement is only likely to be effective in these contexts if the legitimate reasons for absences have been addressed, increasing the likelihood that most doctors will support actions against the free-riding minority. We have described this type of enforcement strategy, which relies on first creating the conditions for successful enforcement as *designing for differences* [16].

Identifying how different doctors vary on their preferences in regard to determinants of absenteeism could inform strategies that would induce most doctors to adhere to rules. While we cannot directly identify doctors who are genuine free riders as opposed to those who may occasionally be absent for good reasons, we can identify those who are unlikely to be persuaded to attend work by any feasible package of incentives, who thus are claiming the benefits of public sector employment without paying any of the costs. While these are the target group for enforcement and sanctions, many more doctors may occasionally be absent but there is scope for feasible changes to incentives that would change their behaviour. Targeted interventions could encourage their compliance with their contractual commitments, leaving a much smaller group who continue to be absent. We hypothesise that this could help create an environment where enforcement against the minority will be supported by the rule-following majority, for example, through peer-monitoring and pressure.

Developing appropriate strategies requires an understanding of the determinants of absenteeism, doctors’ preferences over different components of their job and trade-offs they are willing to make between them. Discrete choice experiments (DCEs) offer a means to do so but we were unable to find any that had examined absenteeism of doctors or other health workers in Bangladesh or elsewhere although some have investigated health worker motivation and retention in other low- and middle-income countries [17]. While context specific, these have highlighted the value that workers place on improved work conditions and benefits, such as bonus payments, promotion, or educational opportunities and show that workers are generally willing to accept certain undesirable job characteristics if balanced by positive ones that they value.

## METHODS

We used a DCE to assess preferences of doctors in regard to determinants of absenteeism in rural facilities in Bangladesh and to identify potential interventions. The project was approved by LSHTM Ethics Committee, Ref. 16248 and Institutional Review Board of BRAC James P Grant School of Public Health, ref. 2017-012. This was conducted by the Anti-Corruption Evidence Consortium (SOAS-ACE) which seeks feasible anti-corruption strategies in Bangladesh, Nigeria and Tanzania. This component forms part of a mixed-methods study examining doctor absenteeism in rural Bangladesh.

### Attribute development

As recommended in the literature [18, 19], DCE attributes were developed through a multi-stage, mixed-methods process including literature review, qualitative interviews and consensus-building workshops with policymakers and health-system stakeholders. The review examined measures to address absenteeism in South and South-East Asia [10]. This informed qualitative interviews with a purposively selected sample of 30 doctors who work or have recently worked in rural facilities, exploring their perceptions of what drives absenteeism and their views on potential solutions. Emerging themes informed attributes for the experiment, which were iteratively refined into nine candidate attributes presented to policymakers and clinicians working in the Bangladeshi health system. Participants in a consensus-building workshop were asked whether the attributes and levels depicted realistic options and their relative policy importance. From this process, six attributes were included in the pilot DCE (Table 1). We theorised that doctors would accept greater monitoring and sanctions against absenteeism if accompanied by greater opportunities or improvements in community relationships, personal security, or rewards for good attendance.

**Table 1.** Candidate attributes included in pilot study and reason for inclusion

| **Attribute** | **Reason for inclusion** | **Propositions** |
| --- | --- | --- |
| **Relationship between the doctor and the local community** | Qualitative work highlighted that the doctors felt that having a good relationship with the community was the key determinant of how manageable the demands placed on them were. Further many noted that if there was a good relationship it would be unlikely that there would be security issues | Doctors expected to prefer facilities that enabled a strong relationship with powerful individuals within the community. |
| **Safety at workplace** | Security was found to be a concern to the doctors both at workplace and living place. Many facilities did not have appropriate or functional security measures while many interviewees told of situations where they had either been verbally or physically abused while in the workplace. | Doctors expected to prefer facilities with effective security measures. |
| **Workload** | Many doctors reported that due to high numbers of patients, staff shortages and social pressures from the local community they found their workload unmanageable and outside of their control. | Doctors expected to prefer facilities with better workload management practices. |
| **Monitoring absenteeism** | Doctors reported that monitoring mechanisms were often either non-functional or inconvenient to use (e.g. having to sign in at central locations away from the facility). | Doctors expected to prefer facilities with either minimal or efficient monitoring systems. |
| **Policies to address absenteeism** | Doctors reported that policies to control absenteeism were often not enforced and findings from the literature suggest there is limited evidence of successful interventions to overcome absenteeism. | Doctors expected to prefer facilities that reward good attendance. Punitive measures were expected to be disliked by most doctors although some noted that appropriate enforcement would reduce absenteeism and thus their personal workload. |
| **Incentive for working in rural facility** | Incentive payments are used in the Bangladesh health system to encourage doctors to work in the Hills districts and would thus be familiar to doctors and allow for a numerical assessment of the importance of attributes. | Doctors expected to prefer higher incentive payments. |

### Piloting

Given this study’s sensitivity and novelty, the questionnaire was extensively piloted over two rounds among 15 doctors at two tertiary hospitals in Dhaka to ensure acceptability, appropriateness, and understandability of the questionnaire. This was followed by short interviews where respondents were asked for feedback on the questions and process. In light of feedback the number of attributes and levels were reduced to the final four outlined in Table 2 and slight changes were made to the way the scenarios were presented.

**Table 2.** Final attributes and levels used in DCE

| **Attribute** | **Level 1** | **Level 2** | **Level 3** | **Level 4** |
| --- | --- | --- | --- | --- |
| **Relationship between the doctor and the local community** | Unsupportive relationship with community | Supportive relationship with community, e.g. monthly meetings of doctors, administrators, community leaders |  |  |
| **Safety at workplace** | No active dedicated security guard and functional CCTV | Active dedicated security guard and functional CCTV |  |  |
| **Career path based on attendance** | Disciplinary action, e.g. salary cut, for poor attendance | Good attendance does not lead to any further benefit. | Good attendance taken into account for promotion and transfer | Good attendance rewarded by bonus points for placement in higher education/training |
| **Incentive for working in rural facility** | No additional financial incentive for posting | Incentive payment of 15% of basic salary for posting | Incentive payment of 30% of basic salary for posting |  |

### DCE design

The hypothetical choice sets for DCE was designed using Ngene software V.1.2.1. We specified a d-efficient, fractional factorial design using a multinomial logit model. No interaction terms were specified in the design. Estimated coefficients for each level were derived from pilot data and used as prior estimates to generate the final survey tool. The final survey consisted of 12 unlabelled choice sets, asking participants to choose between two hypothetical jobs that varied in levels of the attributes outlined in Table 2. A two-stage question format was used to ensure some preference data was collected, while also enabling participants to opt out of accepting the job presented to avoid overestimation of preferences (Figure 1) [20, 21]. Figure 1 shows an example of the choice scenario in English although the final questionnaire was translated into Bengali.

**Figure 1.**
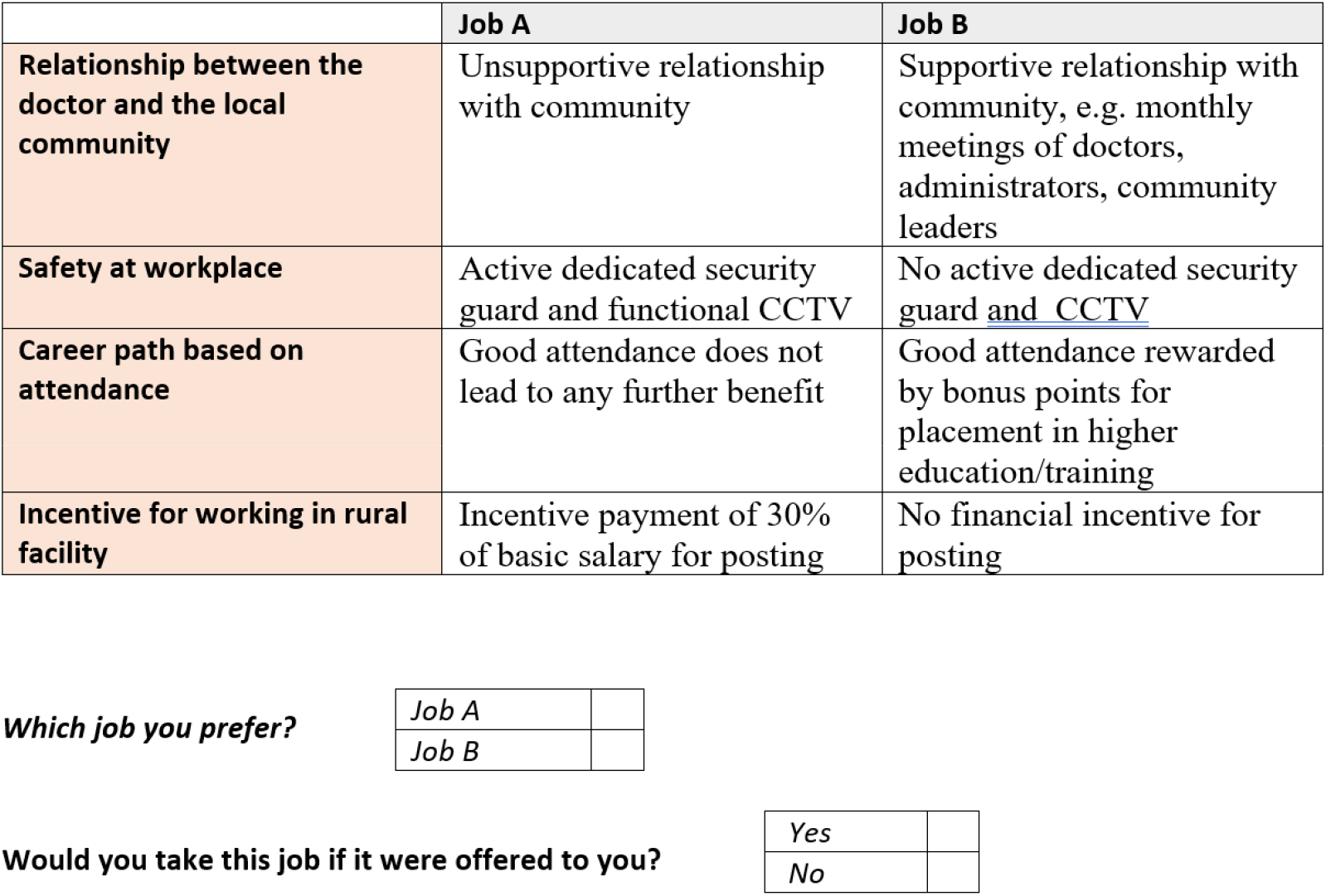
example choice set presented to respondents

### Data collection and sample size

The DCE was conducted in 2019 on doctors currently working at 4 tertiary hospitals in Dhaka with experience of working at rural facilities within the past ten years (mean total rural posting was 3 years, range: 1-12 years). Researchers explained the study, read the introductory statement, explained the job sets, and went through a practice question with all participants. Methods for calculating the required sample sizes for DCEs is contested in the literature [22]. While there is no precise power calculation for DCEs, given our intent to conduct latent class analysis with our data, we targeted a sample size of at least 300 to ensure sufficient power to examine differences across groups [23]. This was in line with previous published DCEs in similar cohorts, including those using latent class analysis [23-26].

### Analysis

DCEs are theoretically based on random utility theory where independent rational actors act to maximise their individual utility [27]; we assume participants chose the job that maximises their individual benefit or utility, which depends on the attributes included in the experiment (Appendix). For the opt-out choice, all attributes were coded as 0. Two models were estimated, the first using panel mixed multinomial logit methods to estimate preferences across all participants, and secondly, latent class analysis to investigate heterogeneity in preferences across our sample. This was to test our hypothesis that incentives will differ significantly for different groups such that some doctors will never accept to work in rural areas. In contrast, others may choose to stay if the right mix of benefits is offered, thus informing interventions targeting this group [11]. Unforced choice data (with options coded as A, B or neither job) was used for all analysis with the respondents’ choices as the dependent variable [21, 28]. All attribute levels were effects coded and, in the mixed model, all parameters were modelled as random with a normal distribution. Constant terms were included to depict respondent preference to not accept either presented job. A three-class model was used in the latent class analysis as model fit statistics showed minimal gains from more classes and class sizes becoming too small for meaningful interpretation (Appendix) [23]. Estimated probabilities were used to assign respondents to groups (with participants assigned to the group with the highest probability of membership) to examine characteristics associated with each group.

A willingness to pay analysis was conducted to estimate the notional amount of incentive payment that respondents would be willing to sacrifice (as a percentage of their base salary) to gain access to different levels of the attributes. Finally, we modelled the potential impact of different policy packages (Table 3) on the probability of accepting a rural job first for the entire sample and then for each identified latent class. All analyses were conducted using NLOGIT software (version 6.0).

**Table 3.** modelled policy scenarios

| Policy Scenario | Supportive community | Presence of security | Disciplinary action for poor attendance | Good attendance considered in promotion and transfer decisions | Good attendance rewarded with bonus points for placement in higher education or training | Incentive payment for posting (per 1% of base salary) |
| --- | --- | --- | --- | --- | --- | --- |
| Baseline | No | No | None | No | No | 0 |
| Punishment only | No | No | Yes | No | No | 0 |
| Punish and reward | No | No | Yes | No | Yes | 0 |
| Punish and support | Yes | Yes | Yes | No | No | 0 |
| Punish, reward and support | Yes | Yes | Yes | No | Yes | 0 |
| Punish and payment | No | No | Yes | No | No | 15% |
| No punish, reward, support and payment | Yes | Yes | No | Yes | Yes | 30% |

### Patient and public involvement

Patients were not involved in this study.

## Results

### General characteristics

The general characteristics of respondents are summarised in Table 3. In total 308 doctors completed the DCE, almost all of whom were married (92%), 46% were female, 76% were aged under 35 and 14% had completed postgraduate training.

### Predictors of choice for the entire sample

The results of the mixed multinomial logit model are presented in Table 4. All attributes significantly predicted the choices of participants. The constant for not accepting either job was the largest predictor of respondents’ choice, suggesting that respondents would only accept a job if it included a suitable set of attributes (across all choices respondents opted-out 41% of the time). The presence of a supportive relationship with the community was the most preferred attribute across the sample, followed by bonus points in the competition for subsequent places in further education or training, consideration of good attendance in promotion and transfer decisions, and personal security. Respondents preferred higher incentive payments but the willingness to pay analysis (Table 4) shows a willingness to sacrifice higher payments in exchange for other positive attributes. The presence of the disciplinary attribute where poor attendance was punished by a salary cut was significantly disliked by respondents. The random parameters’ estimated standard deviation showed significant heterogeneity in preferences for attributes across the sample except for the two attributes relating to rewards for good attendance that appear to be universally preferred.

**Table 4.** Results mixed multinomial logit model for full sample.

| **Table 4 – Results mixed multinomial logit model for full sample** | | McFadden's Pseudo $R^2$ = 0.27, AIC=5960.5, AICc = 5960.6 | | |
| --- | --- | --- | --- | --- |
| **Variable** | **Coefficient** | **Standard error** | **P-value** | **Willingness to pay (as a % payment of base salary)** |
| Constant for not accepting either job | 1.25*** | 0.14 | <0.01 | NA |
| Supportive community | .93*** | 0.05 | <0.01 | 16% (12.4-19.7) |
| Presence of security | .67*** | 0.05 | <0.01 | 12.3% (10.2-14.3) |
| Disciplinary action for poor attendance | -.63*** | 0.06 | <0.01 | -12.9% (-11.8--13.9) |
| Good attendance considered in promotion and transfer decisions | .67*** | 0.06 | <0.01 | 13.5% (12.3-14.7) |
| Good attendance rewarded with bonus points for placement in higher education or training | .77*** | 0.06 | <0.01 | 16 % (14.8-17) |
| Incentive payment for posting (per 1% of base salary) | .05*** | 0.00 | <0.01 | NA |
| **Estimated standard deviations for random parameters** |  |  |  |  |
| Constant for not accepting either job | 2.6*** | 0.16 | <0.01 | NA |
| Supportive community | .6*** | 0.05 | <0.01 | NA |
| Presence of security | .53*** | 0.05 | <0.01 | NA |
| Disciplinary action for poor attendance | .27*** | 0.10 | <0.01 | NA |
| Good attendance considered in promotion and transfer decisions | 0.2* | 0.11 | 0.06 | NA |
| Good attendance rewarded with bonus points for placement in higher education or training | .25* | 0.14 | 0.07 | NA |
| Incentive payment for posting (per 1% of base salary) | .03*** | 0.00 | <0.01 | NA |

### Latent class analysis

The latent class analysis identified three groups with distinct preferences (Table 5), with the most striking difference evident with the constant depicting whether a doctor took a job presented. Group 1 (accounting for 19.8% of our sample) were extremely unlikely to take any job offered, regardless of the attribute levels presented, while Group 2 (32.1%) were unlikely to reject a job presented. Group 3 (48.1%) generally preferred to not accept the job presented, however, the size of this effect was much smaller than for Group 1 and so could be balanced out by the impact of the presented attributes. Estimated background characteristics of latent classes are shown in Appendix.

**Table 5.** Results of latent class analysis

| Variable | Group 1 - 19.8% of sample |  |  | Group 2 - 32.1% of sample |  |  | Group 3 - 48.1% of sample |  |  |
| --- | --- | --- | --- | --- | --- | --- | --- | --- | --- |
| | $\beta$ | Standard error | P-value | $\beta$ | Standard error | P-value | $\beta$ | Standard error | P-value |
| Constant for not accepting either job | 4.47*** | 0.61 | <0.01 | -1.75*** | 0.21 | <0.01 | 1.5*** | 0.11 | <0.01 |
| Supportive community | 1.56*** | 0.38 | <0.01 | .59*** | 0.06 | <0.01 | .8*** | 0.06 | <0.01 |
| Presence of security | .62*** | 0.18 | <0.01 | .43*** | 0.04 | <0.01 | .58*** | 0.05 | <0.01 |
| Disciplinary action for poor attendance | 0.059 | 0.28 | 0.85 | -.50*** | 0.09 | <0.01 | -.61*** | 0.09 | <0.01 |
| Good attendance considered in promotion and transfer decisions | .93*** | 0.33 | 0.01 | .38*** | 0.08 | <0.01 | .64*** | 0.08 | <0.01 |
| Good attendance rewarded with bonus points for placement in higher education or training | -0.07 | 0.49 | 0.8789 | .64*** | 0.09 | <0.01 | .74*** | 0.08 | <0.01 |
| Incentive payment for posting (per 1% of base salary) | .05*** | 0.02 | <0.01 | .04*** | <0.01 | <0.01 | .05*** | <0.01 | <0.01 |

McFaddens' Pseudo $R^2$ = 0.25, AIC=6165.6 AICc = 6331.5
NA – not applicable, \*, \*\*, \*\*\* indicate statistical significance at p=0.1, 0.05 and 0.01 respectively

### Policy simulations

Figure 2 shows how the overall sample and latent groups identified are predicted to respond to different policy packages. Under the baseline scenario, 45.3% of doctors were predicted to accept the job offered, slightly higher than if the only intervention enacted is punishment for poor attendance (44.7%). If punishment for poor attendance is accompanied by a supportive community and secure facility, this proportion grows to 58.7% and 63.9% if good attendance is also rewarded by bonus points for placement in further education. Under the conditions most preferred by doctors, with no punishment accompanied by rewards for good attendance and a 30% incentive payment, 83.4% of doctors would choose to accept a job in a rural facility. While Group 2 were consistently more likely to accept a rural job, the proportions of Groups 1 and 3 were dependent on the specific policy combinations presented. In all but the most attractive scenario, the percentage of doctors in Group 1 accepting a rural posting never rises above 50%.

**Figure 2.**
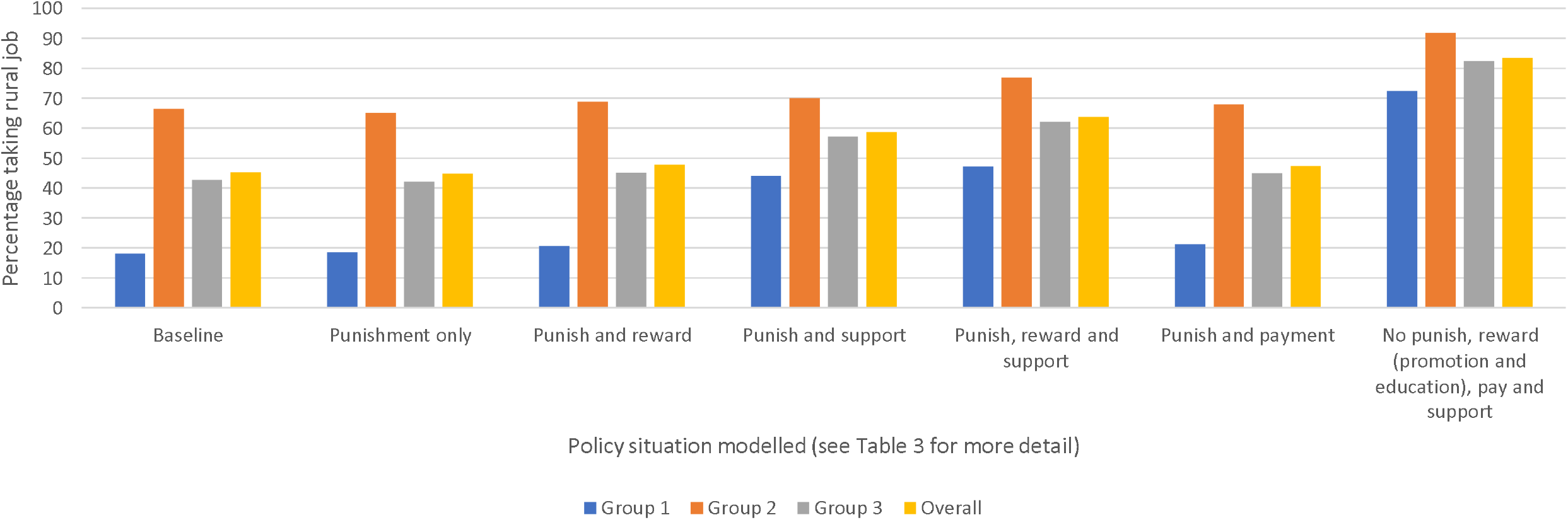
Proportion of respondents who would accept a rural job under different policy scenarios

## Discussion and conclusions

We investigated doctors’ preferences regarding determinants of absenteeism in Bangladesh to inform the development of a package of feasible interventions to overcome this problem. We estimated the potential impact of different interventions on doctors’ behaviour, highlighting the importance of moving beyond traditional regulatory and accountability-focused approaches to addressing absenteeism.[11] Instead, we argued that understanding why doctors leave their posts and what their preferences are, using this information to design targeted interventions to influence their behaviour is potentially more effective. Our findings are broadly consistent with the limited existing literature that has investigated absenteeism but the use of a DCE allows us to assess differences between groups of doctors and so identify a sub-set who appear most amenable to interventions.

From previous literature [11-13], we hypothesised that a significant minority of doctors would be unwilling to accept rural placements even with targeted interventions to improve conditions; this was borne out in our data. Except under the most attractive (and likely costly) conditions, a group of doctors would consistently reject rural posts. Nonetheless, we can now suggest a way of *designing for differences* to achieve enhanced levels of compliance. A substantial minority, almost half the cohort, appears willing to alter their job postings decisions if the right conditions apply. By targeting this group and addressing drivers of ‘reasonable’ absenteeism (such as safety of facilities and fostering better relationships between doctors and the communities that they serve), it appears that attendance can be improved across the system through building a coalition of doctors for whom compliance is in their best interests. We think this will not only have a direct effect on attendance by those responding to these measures, but it will also create an environment in which violations by others are less likely to be tolerated. When levels of compliance increase to levels where doctors who wish to serve their patients can do so without making huge personal sacrifices, a constituency will be created that can exert peer pressure and support the imposition of sanctions on doctors who continue to be absent. Peer pressure is encouraged where individuals believe that those who persist in breaking rules have no good reasons for doing so.

On the other hand, our results are also consistent with historical experience that biometric or other forms of monitoring have not been effective. There are too many doctors engaging in absenteeism and doctors are collectively a powerful group. Large-scale absenteeism also means that peer group pressure is mostly absent as doctors cover each other daily. Interestingly, doctors’ choices in the first latent class were not significantly influenced by the presence of the punishment attribute. This suggests that in addition to being unwilling to attend a rural post, this group had sufficient connections or power to prevent them from being threatened by enforcement mechanisms. Crucially, improving incentives without understanding doctors’ heterogeneity may lead to excessively high levels of incentives (and implemtnation costs) that still fail to work. This crucial finding of our work is novel in the limited literature that has examined health worker absenteeism, which has tended to consider the problem and potential solutions as applying to a uniform set of workers.

While the use of financial incentives is familiar in the Bangladeshi health system as they have been used to encourage workers to take up posts in remote areas like the Chittagong Hill Tracts, feasible salary increases are unlikely to address other key drivers of absenteeism and thus, are unlikely to have a large enough impact to overcome absenteeism sustainably without other concurrent intervention. Our results demonstrate the potential benefits of non-financial incentives, with respondents willing to trade potential incentive payments of significant value in exchange for other job attributes. Many of the interventions we identify are not explicitly focused on absenteeism itself. Still, interventions that increase community support for doctors (often relying on the support of powerful local elites, as shown in the qualitative study) and protect them from the threat of violence by service users, are likely to have many benefits for the health system. Beyond combatting absenteesm, these may help to improve retention and job satisfaction among rural doctors.

Our study has several limitations. First, as with all DCEs, our results are based on respondents’ stated preferences rather than observing actual behaviour. If the stated choices differ from how the doctors would behave in practice our results could be biased. This could be a particular issue in an area as sensitive as absenteeism. We attempted to minimise the impact of such bias through a range of measures including an opt-out option, and an extensive attribute development process followed by piloting. This ensured that the attributes captured the key drivers of absenteeism and job characteristics valued by doctors and were clearly understood by participants. Second, due to resource constraints our sample comprised doctors working in four large urban hospitals with recent experience of working at rural facilities, and thus may not be generaliseable to the broader doctor population. A significant portion of our sample had undertaken postgraduate training already, which could impact the importance placed on the education attribute (if they thought there was no further benefit available to them), however, we sought to make the attribute broad enough to apply to these doctors too and sub-group analysis for these doctors only and with them removed made no significant difference to the results (Appendix).

In conclusion, we were able to suggest a way to address absenteeism, a widespread problem undermining access to and quality of care in LMICs. Top-down regulatory and managerial approaches have had limited success. In contrast, we believe understanding the perspectives of the Bangladeshi doctors about the difficulties they face when working in rural areas and capturing their preferences on different aspects of their jobs can inform pragmatic solutions to reduce absenteeism, which are largely feasible in the context of Bangladesh, and potentially of interest to other LMICs.

## Data Availability

Data collected and held by SOAS-ACE consortium.

## References

1. Adams, A.M., et al., Innovation for universal health coverage in Bangladesh: a call to action. The Lancet, 2013. 382(9910): p. 2104–2111.

2. Gruen, R., et al., Dual job holding practitioners in Bangladesh: an exploration. 2002. 54(2): p. 267–279.

3. Chaudhury, N. and J. Hammer, Ghost doctors: absenteeism in Bangladeshi health facilities. 2003: The World Bank.

4. Joarder, T., et al., Retaining doctors in rural Bangladesh: a policy analysis. 2018. 7(9): p. 847.

5. Rawal, L.B., et al., Developing effective policy strategies to retain health workers in rural Bangladesh: a policy analysis. Hum Resour Health, 2015. 13: p. 36.

6. Ministy of Health and Family Welfare Bangladesh. Office Attendance Monitoring System. 2021 16 February 2021]; Available from: http://103.247.238.92/dghseams/attend/index.php.

7. García-Prado, A. and M. Chawla, The impact of hospital management reforms on absenteeism in Costa Rica. Health Policy and Planning, 2006. 21(2): p. 91–100.

8. Gaitonde, R., et al., Interventions to reduce corruption in the health sector. Cochrane Database of Systematic Reviews, 2016(8).

9. Onwujekwe, O., et al., Corruption in Anglophone West Africa health systems: a systematic review of its different variants and the factors that sustain them. Health Policy and Planning, 2019. 34(7): p. 529–543.

10. Naher, N., et al., The influence of corruption and governance in the delivery of frontline health care services in the public sector: a scoping review of current and future prospects in low and middle-income countries of south and south-east Asia. 2020. 20: p. 1–16.

11. Hutchinson, E., et al., Targeting anticorruption interventions at the front line: developmental governance in health systems. BMJ Global Health, 2020. 5(12): p. e003092.

12. Chaudhury, N. and J.S. Hammer, Ghost doctors: absenteeism in rural Bangladeshi health facilities. The World Bank Economic Review, 2004. 18(3): p. 423–441.

13. Naher, N., et al., A study of absenteeism among doctors in rural Bangladesh. 2020.

14. Khan, M., Political settlements and the analysis of institutions. African Affairs, 2018. 117(469): p. 636–655.

15. Behuria, B., Buur, L, Gray, H,, Studying political settlements in Africa. African Affairs, 2017. 116(464): p. 508–525.

16. Khan, M., P. Roy, and A. Andreoni, Anti-corruption in adverse contexts: strategies for improving implementation. 2019.

17. Mandeville, K.L., M. Lagarde, and K.J.B.h.s.r. Hanson, The use of discrete choice experiments to inform health workforce policy: a systematic review. 2014. 14(1): p. 367.

18. Lancsar, E. and J.J.P. Louviere, Conducting discrete choice experiments to inform healthcare decision making. 2008. 26(8): p. 661–677.

19. Coast, J., et al., Using qualitative methods for attribute development for discrete choice experiments: issues and recommendations. 2012. 21(6): p. 730–741.

20. Mandeville, K.L., M. Lagarde, and K. Hanson, The use of discrete choice experiments to inform health workforce policy: a systematic review. BMC health services research, 2014. 14(1): p. 1–14.

21. Scott, A., et al., Getting doctors into the bush: General Practitioners’ preferences for rural location. Social Science & Medicine, 2013. 96: p. 33–44.

22. de Bekker-Grob, E.W., et al., Sample size requirements for discrete-choice experiments in healthcare: a practical guide. 2015. 8(5): p. 373–384.

23. Zhou, M., W.M. Thayer, and J.F. Bridges, Using latent class analysis to model preference heterogeneity in health: a systematic review. Pharmacoeconomics, 2018. 36(2): p. 175–187.

24. Abdel-All, M., et al., What do community health workers want? Findings of a discrete choice experiment among Accredited Social Health Activists (ASHAs) in India. 2019. 4(3): p. e001509.

25. Kruk, M.E., et al., Rural practice preferences among medical students in Ghana: a discrete choice experiment. 2010. 88: p. 333–341.

26. Blaauw, D., et al., Policy interventions that attract nurses to rural areas: a multicountry discrete choice experiment. 2010. 88: p. 350–356.

27. Hensher, D.A. and L.W. Johnson, Applied discrete-choice modelling. 2018: Routledge.

28. Abdel-All, M., et al., What do community health workers want? Findings of a discrete choice experiment among Accredited Social Health Activists (ASHAs) in India. BMJ Global Health, 2019. 4(3): p. e001509.

