## Appendix for "How to incentivise doctor attendance in Bangladesh: a latent class analysis of a discrete choice experiment"

Supplementary Appendix

Theoretical background to analysis

DCEs are theoretically based on random utility theory where independent rational actors act to maximise their individual utility [28]. We assume participants will choose the job that maximises their individual benefit or utility which depends on the attributes such that:

$$U \left( Job A or Job B \right)= \beta_{1}{*Relationship+ \beta}_{2}*Safety+\beta_{3}*{Punishment+\beta}_{4}*Promotion+\beta_{5}*Education*\beta_{6}* Incentive+ \varepsilon$$

Where:

Relationship = The relationship with the local community;

Safety = Whether there were active security guards and CCTV;

Punishment = The presence of disciplinary action for poor attendance;

Promotion = Good attendance considered in promotion and transfer decisions;

Education = Good attendance rewarded by bonus points for placement in further education;

Incentive = Incentive payment attached to the post.

For the opt-out choice, all attributes were coded as 0 such that $U\left( No job taken \right)=0$.

Model fit statistics for latent class models

| **Table S1 - Number of latent classes** | **2** | **3** | **4** | **5** |
| --- | --- | --- | --- | --- |
| Log-likelihood function | -3198.77 | -3059.78 | -3010.29 | -2972.52 |
| Pseudo R^2 | 0.212 | 0.246 | 0.259 | 0.268 |
| AIC | 6427.5 | 6165.6 | 6082.6 | 6023 |
| AICc | 6535.8 | 6331.5 | 6306.2 | 6304.4 |
| BIC | 6520.8 | 6308.5 | 6275.2 | 6265.4 |
| Size of the smallest group (proportion of sample) | 39.9% | 20.8% | 13.9% | 7.5% |
| Size of the smallest group (estimated respondents) | 123 | 64 | 43 | 23 |

AIC: Akaike information criterion

AICc: Akaike information criterion with a correction for finite sample sizes

BIC: Bayesian information criterion

| **Table S2 – General characteristics of estimated groups** | Group 1 | Group 2 | Group 3 |
| --- | --- | --- | --- |
| Average age | 34.2 | 33.2 | 33.1 |
| Proportion aged over 40 | 10.77% | 7.84% | 11.35% |
| Proportion female | 47.69% | 50.00% | 43.26% |
| Completed Postgraduate training | 15.38% | 14.71% | 12.77% |
| Proportion with 2 or more kids | 47.69% | 48.04% | 33.33% |
| Proportion who rated their financial situation over the past year as good or very good | 63.08% | 54.90% | 51.06% |
| Proportion who responded professional network was important to promotion | 32.31% | 32.35% | 28.37% |
| Proportion who responded personal network was important to promotion | 20.00% | 18.63% | 26.95% |
| Proportion who responded political network was important to promotion | 32.31% | 32.35% | 36.88% |
| Proportion that experienced any challenge in their previous rural post | 98.46% | 98.04% | 100.00% |
| Proportion member of a professional association | 71.88% | 68.32% | 79.14% |
| Proportion who served their full rural placement without interruption | 80.00% | 67.00% | 76.81% |

Estimated characteristics of latent class groups

Mixed multinomial results excluding respondents with postgraduate training

| **Table S3 – Results mixed multinomial logit model for sample excluding those with postgraduate education** | McFadden’s Pseudo R^2^ = 0.27 | | |
| --- | --- | --- | --- |
| **Variable** | **Coefficient** | **Standard error** | **P-value** |
| Constant for not accepting either job | 1.28*** | 0.16 | <0.01 |
| Supportive community | .97*** | 0.06 | <0.01 |
| Presence of security | .67*** | 0.05 | <0.01 |
| Disciplinary action for poor attendance | -.64*** | 0.07 | <0.01 |
| Good attendance considered in promotion and transfer decisions | .67*** | 0.06 | <0.01 |
| Good attendance rewarded with bonus points for placement in higher education or training | .82*** | 0.07 | <0.01 |
| Incentive payment for posting (per 1% of base salary) | .06*** | <0.01 | <0.01 |
| **Estimated standard deviations for random parameters** |  |  |  |
| Constant for not accepting either job | 2.6*** | 0.17 | <0.01 |
| Supportive community | .64*** | 0.06 | <0.01 |
| Presence of security | .56*** | 0.05 | <0.01 |
| Disciplinary action for poor attendance | .34*** | 0.10 | <0.01 |
| Good attendance considered in promotion and transfer decisions | 0.3*** | 0.10 | <0.01 |
| Good attendance rewarded with bonus points for placement in higher education or training | .17 | 0.11 | 0.11 |
| Incentive payment for posting (per 1% of base salary) | .03*** | <0.01 | <0.01 |
